# Forecasting hospitalizations due to COVID-19 in South Dakota, USA

**DOI:** 10.1101/2020.07.22.20160184

**Authors:** Jeff S. Wesner, Dan Van Peursem, José D. Flores, Yuhlong Lio, Chelsea A. Wesner, University of South Dakota

## Abstract

Anticipating the number of hospital beds needed for patients with COVID-19 remains a challenge. Early efforts to predict hospital bed needs focused on deriving predictions from SIR models, largely at the level of countries, provinces, or states. In the United States, these models rely on data reported by state health agencies. However, predictive disease and hospitalization dynamics at the state level are complicated by geographic variation in disease parameters. In addition it is difficult to make forecasts early in a pandemic due to minimal data. However, Bayesian approaches that allow models to be specified with informed prior information from areas that have already completed a disease curve can serve as prior estimates for areas that are beginning their curve. Here, a Bayesian non-linear regression (Weibull function) was used to forecast cumulative and active COVID-19 hospitalizations for South Dakota, USA. As expected, early forecasts were dominated by prior information, which was derived from New York City. Importantly, hospitalization trends also differed within South Dakota due to early peaks in an urban area, followed by later peaks in other rural areas of the state. Combining these trends led to altered forecasts with relevant policy implications.

## 2 Introduction

The novel coronavirus (SARS-CoV-2) was first detected in December 2019 in Wuhan, China and has since spread globally. The disease caused by SARS-CoV-2 (COVID-19) can lead to hospitalization or death in all age groups, but particularly in older age groups with comorbidities such as hypertension, obesity, and diabetes.^1^ For example, in France, Salja et al.^2^ estimated that 3.6% of infected people become hospitalized, but that rate varies from a low of 0.001% to 10.1% for individuals that are <20 years old versus those >80 years old, respectively. A central challenge for hospitals is predicting how many hospitalizations will occur due to COVID-19, and whether hospital capacities will be exceeded.

Predicting hospitalization needs due to COVID-19 may be particularly challenging in rural areas. For example, relative to urban areas, rural communities in the U.S. have reduced access to health care^3^ and increased mortality from chronic diseases,^4^ both of which are key risk factors for COVID-19. South Dakota is among the most rural states in the U.S. with a 2017 population estimate of 869,666.^5^ Of South Dakota’s 66 counties, 52% are frontier (with <15.5 persons per square kilometer) and 32% encompass or are comprised of reservation lands representing nine federally recognized tribes. Communites in rural areas also tend to have older populations. For example, among 56 U.S. counties with the largest proportion of people ages >85, all but two counties are rural and a county in South Dakota ranks first.^6^ South Dakota’s public health infrastructure is limited with a centralized state department of health that delivers public health services through a network of regional and county offices, most of which house a single public health nurse. Nearly 80% of nonprofit hospitals in South Dakota are critical access hospitals with 25 or fewer acute inpatient beds,^7^ and acess to medical facilities with an intensive care unit (ICU) and ventilators is limited in rural areas.^8^

To our knowledge, there are no published studies that model hospitalizations due to COVID-19 in rural and low resource settings. Developing new ways to model infectious disease outbreaks in jurisdictions with limited public health and health care infrastructure is critical to preventing and reducing mortality and morbidity in communities that are at high risk of COVID-19. Early predictions of hospitalization in the United States relied on projections from SIR models and their derivatives.^9^ Because they are developed primarily to simulate disease spread through a hypothetical, well-mixed population, SIR-based models require a number of assumptions to generate predictions of hospitalizations or death.^10^ Unlike many U.S. states, South Dakota has publically released hospitalization data daily and there is now enough data to model hospitalization curves directly, rather than infering hospitalizations through an SIR.

Here, we modeled cumulative hospitalizations in an urban (Minnehaha) versus rural population within South Dakota using a Bayesian non-linear Weibull function. Because early predictions in a disease outbreak are critical for planning, but also are data limited, we used informed priors from New York City, which began its hospitalization curve before South Dakota. While New York City is not a rural area, the use of informed prior distributions allowed our model to make reasonable, though highly uncertain, forecasts of hospitalizations in a rural setting.

## 3 Methods

### Data Retrieval

We obtained data on cumulative hospitalizations and active hospitalizations (number hospitalized on a given day) from the data dashboard for the South Dakota Department of Health (SD DOH) - https://doh.sd.gov/news/Coronavirus.aspx#SD. Data for cumulative hospitalizations began on 2020-03-08 and were entered by hand into a .csv each day (SD DOH only reports totals for the current day, not a timeline). Data for active hospitalizations were not released until 2020-04-20, when 56 people were actively hospitalized. Beginning on that date, we also updated our .csv with active hospitalizations each day.

When data collection began, most cases and hospitalizations were located in Minnehaha County, South Dakota. Therefore, during data collection, we noted hospitalizations in Minnehaha County versus the rest of South Dakota to capture any potential divergent trends under the assumption that the disease would spread more slowly across rural South Dakota. For these two areas, data are only available for cumulative hospitalizations, not for active hospitalizations. Minnehaha County has a population density of 619 people per *km*^2^, which is >50 times higher than the state average population density of 28 people per *km*^2^.^5^ Comparing these two areas allowed us to model COVID-19 hospitalizations in a rural and urban setting within the same state.

### Models

We estimated cumulative hospitalizations using a Bayesian model in which hospitalizations were modeled as a sigmoid function of time using the Weibull function.^11,12^ The Weibull function is derived from the Weibull cumulative distribution^13^ and has been used widely in biology to model growth curves.^14^ We chose the Weibull function because it is more flexible than the logistic function and is asymmetric around the inflection point.^11,15^ We fit the Weibull function to two sets of data that describe 1) the cumulative hospitalizations for the state of South Dakota and 2) the cumulative hospitalizations for subgroups of Minnehaha County and the rest of South Dakota. Because the data were counts with positive outcomes, we used a Poisson likelihood with a log-link.

#### Model 1

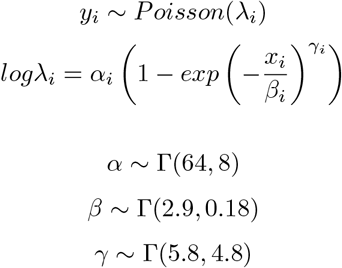

With the above notation, *y*_*i*_ is the cumulative number of people hospitalized in South Dakota on the *i*^*th*^ date, *α* is the asymptote, *β*is the inflection point, and *γ* is the slope at the inflection point. Gamma priors were used because each parameter must be positive and continuous.

Informative prior distributions were derived from the cumulative hospitalization curve in New York City (NYC Department of Health, https://www1.nyc.gov/site/doh/covid/covid-19-data.page). We derived the priors from New York City because NYC had nearly completed its hospitalization curve when South Dakota’s was still beginning and because the data were available as a timeline (many states either have not reported temporal hospitalization data or have not made the data easily extractable).

To derive prior distributions for South Dakota, we first fit the aforementioned model to NYC’s hospitalization curve. Before fitting t he m odel, w e m ultiplied N YC h ospitalizations b y 0 .10 t o p ut t hem o n t he s cale of South Dakota’s population (which is ∼10% of NYC’s population). We then fit t he model to t hese adjusted hospitalizations using prior values of Γ(1.2, 0.1) for *α*, Γ(0.25, 0.005) for *β*, and Γ(1.4, 0.3) for *γ*. Those reflect p rior d istributions w ith w ide s tandard deviations t hat would represent a p otential overload of South Dakota’s ∼ 2000 hospital beds: 10,000 +/-5000 (mean +/-sd) for *α*, 50 +/-100 for *β*, and 100 *±* 50 for *γ*.

#### Model 2

To capture trends inside and outside of Minnehaha County, we fit the same model as before, but included an indicator variable with two levels (Minnehaha County or Outside Minnehaha County) for each of the three parameters.

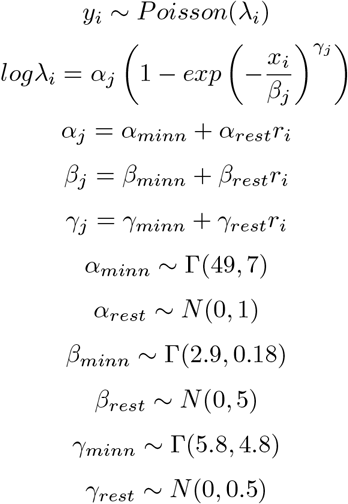

With the above notation, *y*_*i*_ is the cumulative number of people hospitalized in each *i* date (*x*_*i*_), *α*_*j*_, *β*_*j*_, and *γ*_*j*_ are the parameters for each *j* group (Minnehaha County or the Rest of South Dakota), *X*_*minn*_ are the priors for each X parameter (*α, β, γ*), *X*_*rest*_ are the priors for the difference in parameter values between Minnehaha and the Rest of South Dakota, and *r*_*i*_ is an indicator variable that is 0 if the data are in Minnehaha County and 1 otherwise.

As before, prior values were chosen from a combination of prior information from NYC and from prior predictive simulation.^16^ To do this, we simulated 300 cumulative hospitalization curves with mean values for each parameter derived from the fit of the NYC model. Because NYC has both a higher absolute population size and a higher population density (by 10-fold) than South Dakota, we adjusted prior means and standard deviations so that the prior predictive distributions estimated hospitalizations to have a maximum that is slightly below the maximum of NYC, but with standard deviations that still include positive prior probability for some extreme prediction (e.g., 50,000 cumulative hospitalizations). Figure 1 shows the prior predictions for both models.

##### Markov Chain Monte Carlo

Each model aforementioned was specified in R (version 3.6.3; R Core Team 2020) using the *brms* package.^17^ Posterior sampling was performed using Hamiltonian Monte Carlo in *rstan* (version 2.19.2,^18^). We fit four chains, each with 2000 iterations, discarding the first 1000 iterations of each chain as warm-up. Warm-up samples are similar to burn-in sampling, but are used in this case as an optimizer for the HMC algorithm. Chains were checked for convergence using trace plots to assure overlap (*Supplementary Information*), and by ensuring that the Gelman-Rubin convergence diagnostic 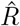 was < 1.1.^19^

##### Posterior prediction

To forecast cumulative hospitalizations, we used the posterior predictive distributions from each model by first solving for the fitted values across each iteration of the posterior:

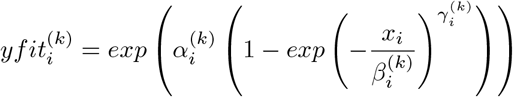

where *k* is the *k*^*th*^ iteration from the posterior distribution and *i* is the *i*^*th*^ date. Posterior predicted values were estimated by drawing each 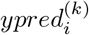 from the Poisson distribution:

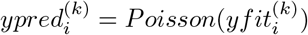

We then summarized the mean, median, standard deviation and credible intervals (50 and 95%) across the posterior distribution of fitted and predicted values. For visualization, we plotted fitted values within the range of the data and predicted values beyond the range of the data.

##### Estimating Active Hospitalizations

To estimate active hospitalizations from the cumulative hospitalization curve, we first derived daily incidence *ϕ* for each iteration of 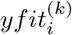 in which

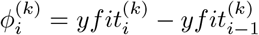

We then summed incidence over the previous 5, 10, 12, or 15 days to estimate variable lengths of hospital stays:

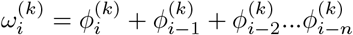

where 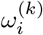 is the number of people actively hospitalized on the *i*^*th*^ day for the *k*^*th*^ iteration, 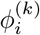 is the incidence on the *i*^*th*^ day for the *k*^*th*^ iteration and *n* is 5, 10, 12, or 15. These lengths of stay were chosen to capture the range of reported hopsital stay lengths from the literature.^20^

We then plotted these predictions against active hospitalizations reported by the South Dakota Department of Health. For the group levels (*Model 2*), we performed the same calculations as above, but for each group. In addition, we also estimated the state-level hospitalizations from *Model 2* by summing the predictions from each group. This allowed us to compare predictions when only state-level data were available versus predictions with data available for different areas of the state.

##### Parameter change over time

We re-fit the model each day as data were released. To visualize how parameter values changed over time as data were added, we plotted posterior predictions and parameter values from model runs in 20 day intervals. Twenty days was arbitrarily chosen to allow for visual clarity in the plots.

## 4 Results

At the state level, model 1 predicted a total of 932 hospitalizations (median) in South Dakota (90% CrI: 808-977, Table 2). The inflection point was predicted at 38 days after the first hospitalization, suggesting that the peak rate of hospitalizations occurred around April 20, 2020 (Table 2). In contrast, the model with group-level effects clearly showed that hospitalizations trends differed in Minnehaha County verses the rest of South Dakota (Figure 2). In Minnehaha County, the inflection point occurred around day 23 and revealed an asymptote of 332 hospitalizations (90% CrI: 326-338) (Table 2). In the rest of South Dakota, the inflection point occurred ∼40 days later (day 62, Figure 2). Similarly, the maximum cumulative hospitalizations in the rest of South Dakota are estimated at a median of ∼811, but with large uncertainty (90% CrI: 745-891, Table 2).

**Figure 2:**
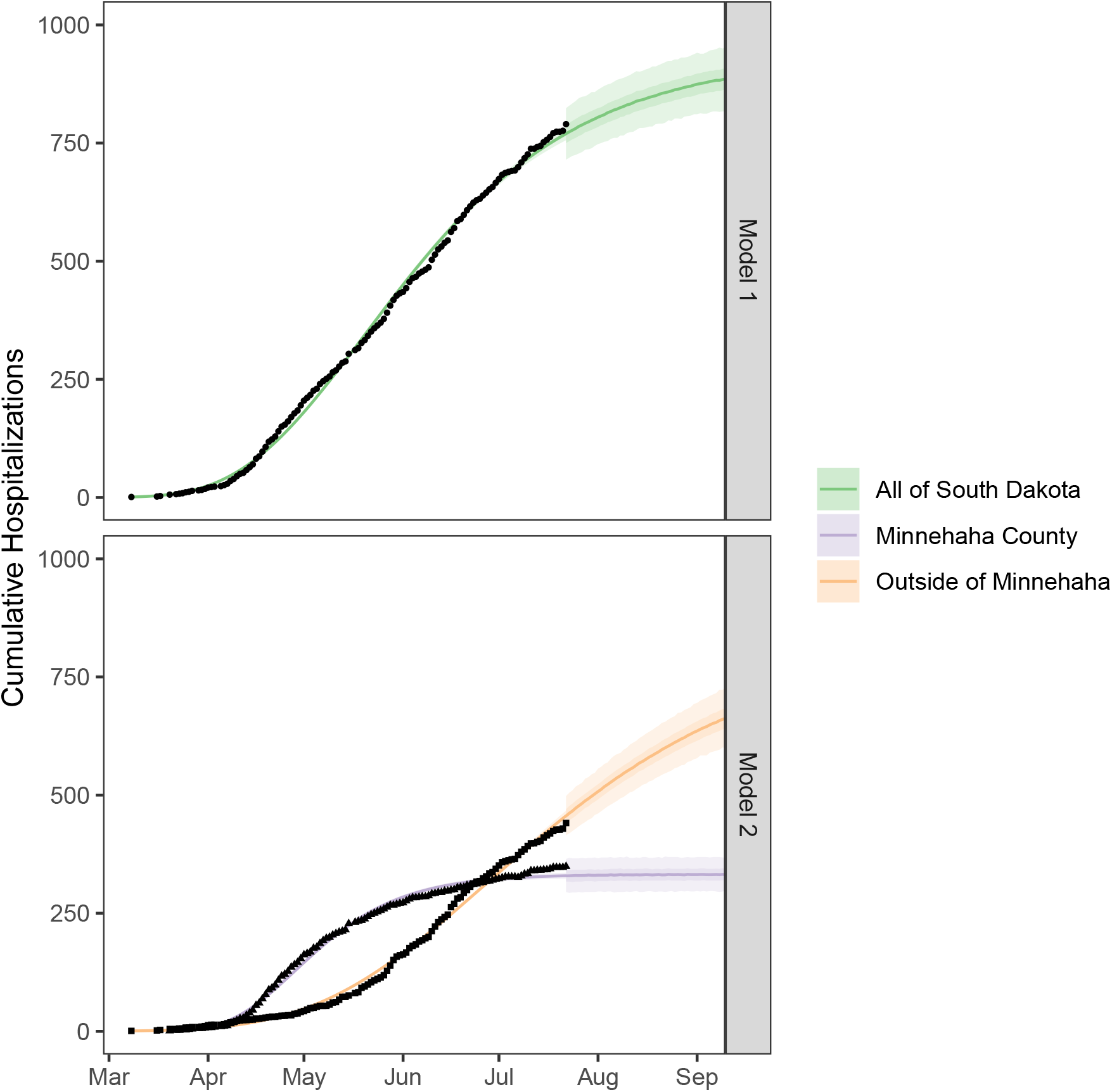
Posterior distributions of cumulative hospitalizations in South Dakota. Lines indicate medians and shading indicates the 50 and 90% intervals. Predictions beyond the data represent samples from the posterior predictive distribution. Predictions within the data represent samples from the posterior fitted distribution.

**Table 1:** Posterior distributions from the New York City model and prior distributions for the South Dakota models.

| Parameter | Model | Post_Prior | Mean | SD | Shape | Rate |
| --- | --- | --- | --- | --- | --- | --- |
| $\alpha$ | nyc | posterior | 8.58 | 0.00 | 12576784.38 | 1465706.21 |
| $\alpha$ | m1 | prior | 7.98 | 0.98 | 65.83 | 8.25 |
| $\alpha$ | m2 | prior | 7.02 | 1.01 | 48.12 | 6.86 |
| $\alpha_{rest}$ | m2 | prior | 0.00 | 1.00 | NA | NA |
| $\beta$ | nyc | posterior | 17.65 | 0.07 | 66915.06 | 3791.95 |
| $\beta$ | m1 | prior | 16.50 | 10.20 | 2.62 | 0.16 |
| $\beta$ | m2 | prior | 16.86 | 9.84 | 2.94 | 0.17 |
| $\beta_{rest}$ | m2 | prior | 0.00 | 5.00 | NA | NA |
| $\gamma$ | nyc | posterior | 1.26 | 0.01 | 23182.63 | 18340.23 |
| $\gamma$ | m1 | prior | 1.21 | 0.47 | 6.64 | 5.48 |
| $\gamma$ | m2 | prior | 1.20 | 0.53 | 5.21 | 4.33 |
| $\gamma_{rest}$ | m2 | prior | 0.00 | 0.50 | NA | NA |

**Table 2:** Summary statistics of model parameters. Asymptotes are exponentiated to place them on the scale of the response variable (cumulative hospitalizations). Summaries are derived from the posterior distributions of each parameter in the corresponding model.

| Model | Group | Parameter | Median | Mean | SD | Low90 | Upper90 |
| --- | --- | --- | --- | --- | --- | --- | --- |
| Model 1 | South Dakota | $\beta$ | 37.8 | 37.8 | 0.3 | 37.3 | 38.3 |
| Model 1 | South Dakota | $\exp(\alpha)$ | 932.9 | 934.7 | 24.3 | 898.3 | 977.7 |
| Model 1 | South Dakota | $\gamma$ | 1.0 | 1.0 | 0.0 | 1.0 | 1.0 |
| Model 2 | Minnehaha | $\beta_{minn}$ | 23.1 | 23.1 | 0.3 | 22.6 | 23.6 |
| Model 2 | Minnehaha | $\exp(\alpha_{minn})$ | 332.1 | 332.2 | 3.7 | 326.3 | 338.4 |
| Model 2 | Minnehaha | $\gamma_{minn}$ | 1.1 | 1.1 | 0.0 | 1.1 | 1.2 |
| Model 2 | Outside of Minnehaha | $\beta_{minn} + \beta_{rest}$ | 61.6 | 61.7 | 0.9 | 60.3 | 63.2 |
| Model 2 | Outside of Minnehaha | $\exp(\alpha_{minn} + \alpha_{rest})$ | 811.9 | 814.5 | 44.9 | 745.2 | 891.3 |
| Model 2 | Outside of Minnehaha | $\gamma_{minn} + \gamma_{rest}$ | 1.1 | 1.1 | 0.0 | 1.1 | 1.2 |

As expected the uncertainty in predictions (Figure 3) improved over time as data were added to the model. The largest improvement appeared to occur during the model run on day 60, when all parameter values appeared to stabilize (*Supplementary Information*). After this date, predictions for hospitalizations in Minnehaha County were stable, while predictions outside of Minnehaha County tended to underpredict future hospitalizations until the most recent model runs (Figure 3).

**Figure 3:**
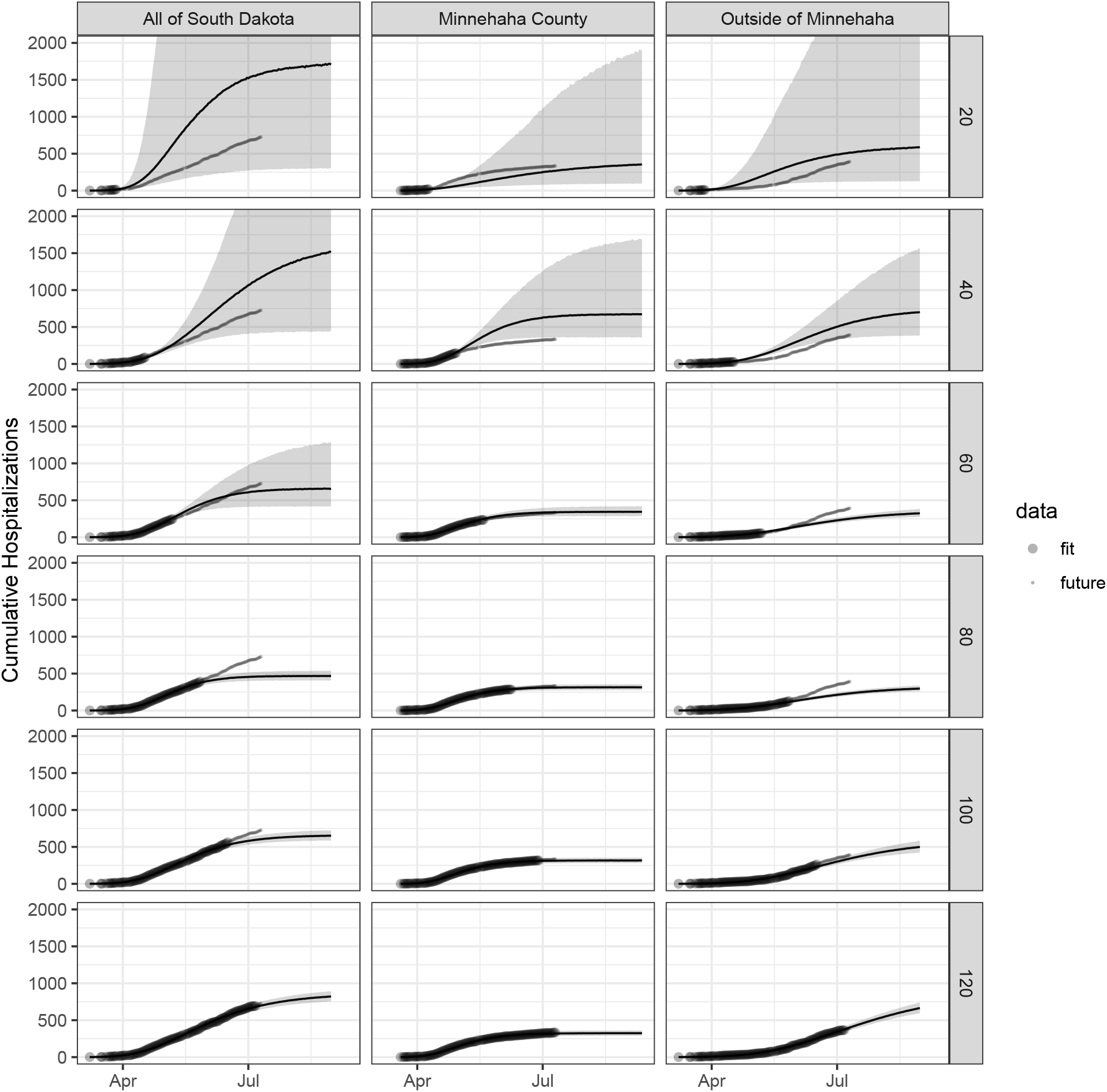
Change in predictions over time as models are fit using data at days 20, 40, 60, 80, 100, and 120. Data points show the full hospitalization curve (same for each panel). The size of the data points changes based on what data were used to fit the model versus the actual data obtained after a given model was fit.

Converting the cumulative curve to estimate the number of people actively hospitalized yields a maximum estimate of ∼100 people actively hospitalized. This estimate is derived from the assumption that an average patient will spend 10 days in the hospital. That assumption appeared to best approximate the state reported data best (Figure 4). One clear difference between the two models is that the state-level model predicts a single peak in active hospitalizations, while model 2 predictions two distinct peaks (Figure 4). The data appear consistent with two separate peaks. However, active hospitalization data are only available for the state, so we are unable to compare data to predictions at the group-level (Figure 4).

**Figure 4:**
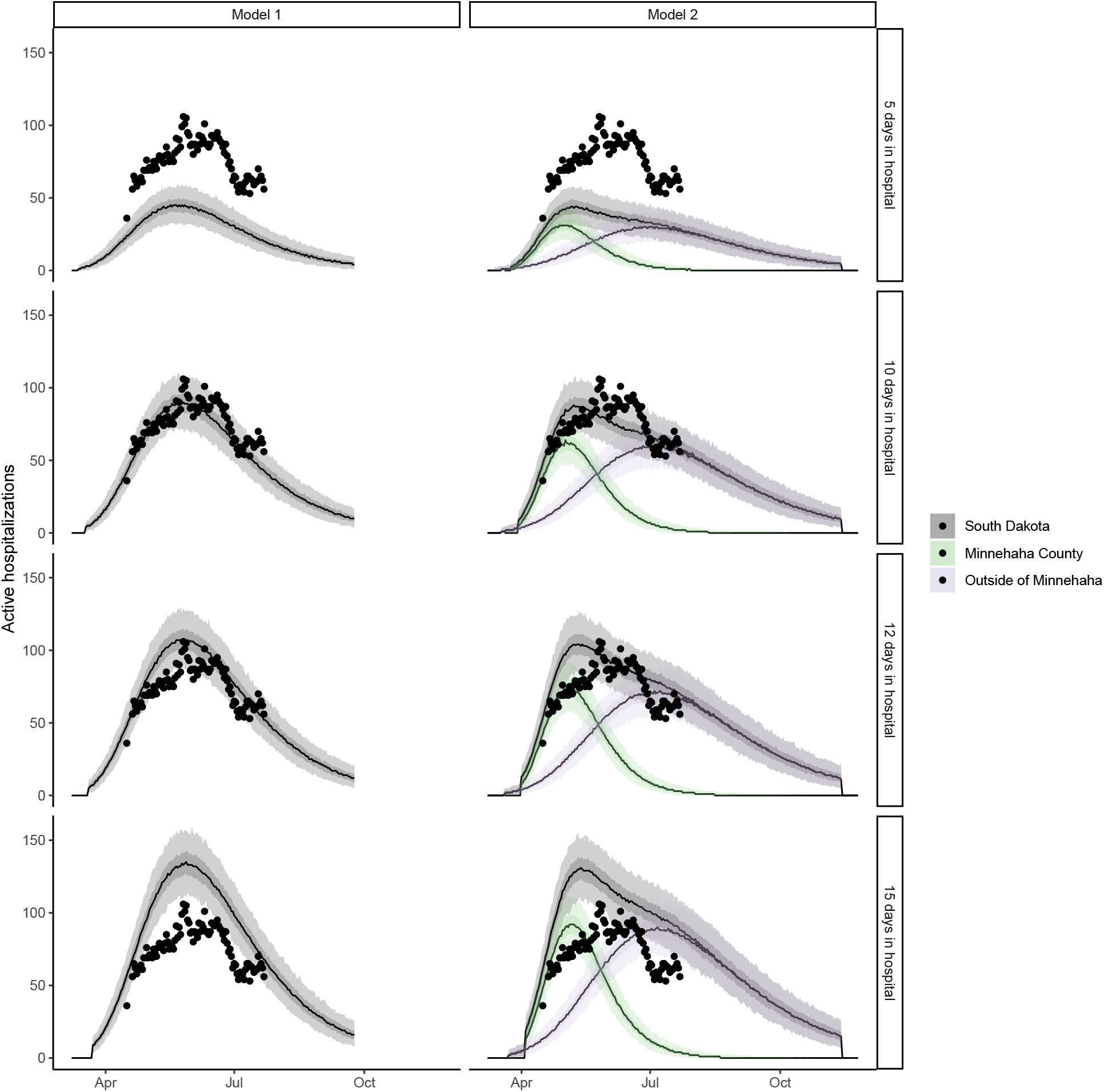
Posterior predictive distributions of active hospitalizations in South Dakota. Lines indicate medians and shading indicates the 50 and 95% prediction intervals.

## 5 Discussion

The most important result of this study is that modeling trends separately in urban versus rural parts of a state population reveal different projections of cumulative hospitalizations than if modeled only using state-level data. In particular, the model with urban vs. rural groups predicts that Minnehaha County will attain a maximum of ∼316 hospitalizations, while areas outside of Minnehaha will attain a maximum of ∼785. That results in approximately ∼1000 people cumulatively hospitalized. In contrast, the model with only state-level data indicates a 24% smaller number of total people hospitalized (762).

In addition to differences in maximum hospitalizations, the models predict different trends of active hospitalizations. Active hospitalizations are more relevant than cumulative hospitalizations for measuring hospital capacity. They have been modeled for the COVID pandemic in a variety of ways with many states adopting a version of an SIR model, such as the CHIME model.^9^ One challenge with estimating hospitalizations using an SIR-based approach is that they require estimates of a number of transition probabilities to convert infections into hospitalizations. These include estimating the proportion of infected people that become symptomatic, the proportion of those that require hospitalization, the proportion of those that seek hospitalization, the time lag between viral infection as modeled in an SIR, symptom onset, and actual hospitalization.^9,10^ Early in an epidemic, when hospitalization data are scarce or absent, these models are essential for predicting possible hospitalization scenarios. However, once enough data are available on hospitalizations, it is possible to model the hospitalization curve directly, as we have done here.

Though we did not attempt to predict COVID-19 infections in South Dakota, it may be possible to use our approach to do so by treating infections as a latent variable that leads to subsequent hospitalizations. Flaxman et al. ^10^ provide an example of this approach using COVID-19-related deaths rather than hospitalizations. By modeling the dynamics of deaths, they estimated a time-varying reproduction number (*R*_*t*_), which was used to estimate the number of positive cases. Using deaths (or hospitalizations) to estimate infection dynamics may help to overcome limitations in testing capacity, which in turn lead to difficulty in linking publically-reported testing results to true population-level infection rates. This is particularly true in the United States, which has limited testing capacity and no centrally coordinated testing program.^21^

An advantage to the Bayesian approach is that we can use prior values of parameters in a model to fit a model with limited data. In our case, those parameters were conveniently available from New York City. However, if we were modeling a hospitalizaton curve that had no prior estimates, then we might derive priors from another epidemic that had similar disease characteristics or use prior predictive simulation to bound the model to reasonable prior predictions^16^. In particular, because the prior distributions for individual parameters may not be known or are difficult to interpret without the consideration of the likelihood,^22^ it is important to assess the implications of prior choices using the prior predictive distribution (i.e. simulating potential from the prior distrbutions alone)^16^ (Figure 1). In our model, data simulated from the prior helped to confirm that our model was specified in a way that included a wide range of hospitalization trajectories (Figure 1), but excluded extreme values that might have come from more diffuse priors, such as projecting asymptotes with hospitalizations that are higher than the population of South Dakota.

**Figure 1:**
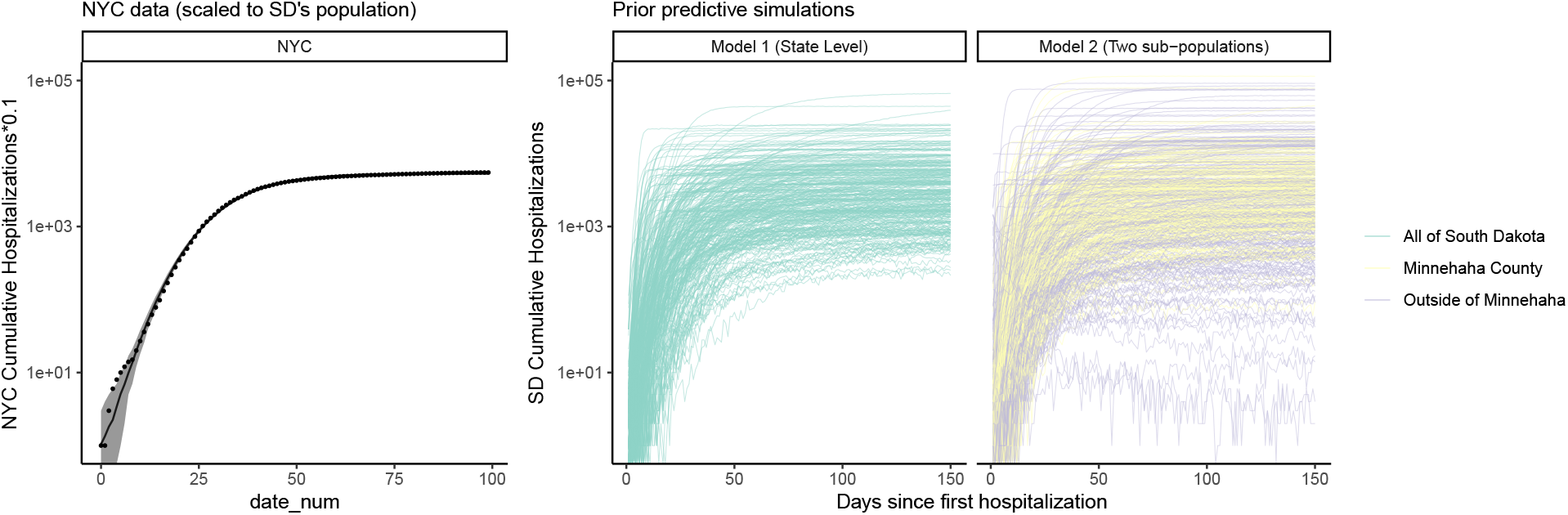
Left: Model of New York City’s hospitalization curve. Data are divided by 10 to reflect the relative population sizes in South Dakota versus New York city. Right: Three-hundred simulations of cumulative hospitalizations from the prior predictive distribution of each model for South Dakota. Priors for Model 1 were derived from the fit of NYC’s hospitalization curve. Priors for Model 2 were similar to those of Model 1, but had a reduced prediction of cumulative hospitalizations to account for the smaller populations of each group (Minnehaha County vs Outside Minnehaha County) relative to the whole state population.

## Data Availability

All data and code are available at https://github.com/jswesner/covid_sd_ms. These will be permanently archived via Zenodo upon acceptance of this manuscript.

## 6 Data Availability Statement

All data and code are available at https://github.com/jswesner/covid_sd_ms.^23^ These will be permanently archived via Zenodo upon acceptance of this manuscript.

## 7 Acknowledgments

We declare no conflicts of interest. We thank the South Dakota Department of Health for making the hospitalization data publically available. This work is not affiliated with any funding agency.

## 11 Supplementary Information

Here we compare the prior and posterior distributions of each parameter in the models. Figure S1 indicates how much information was learned about each parameter from adding data. Trace plots indicate well-mixed chains for each model.

**Figure 5:**
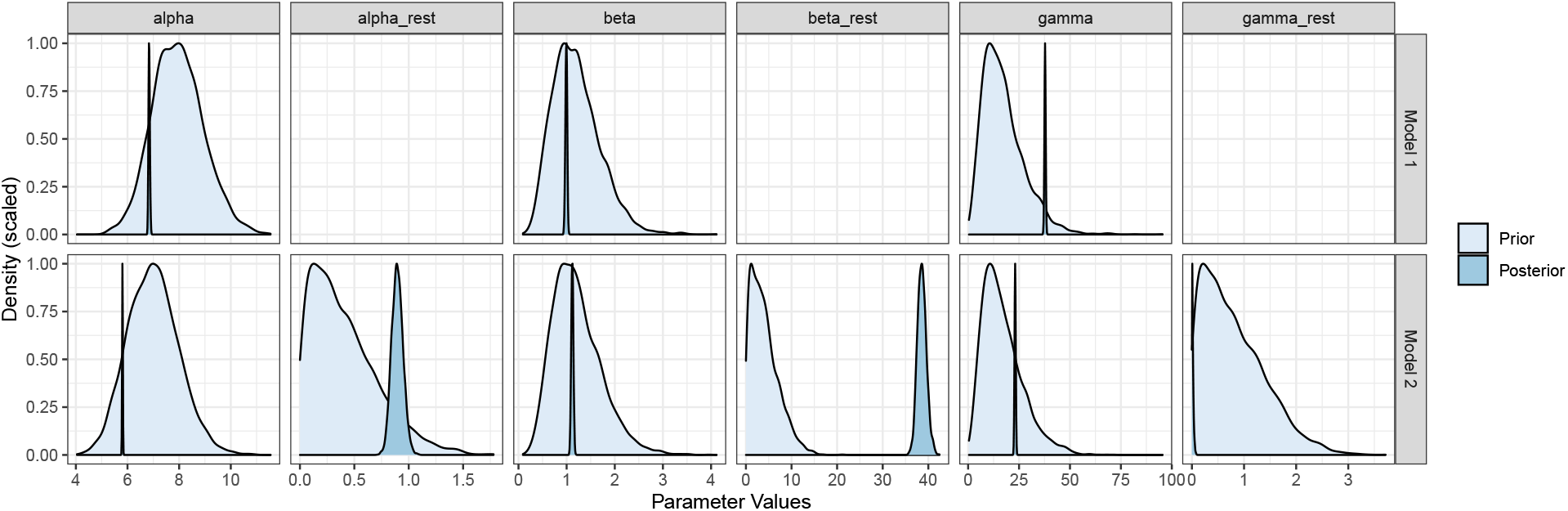
Comparison of prior and posterior distributions for each parameter in models 1 and 2.

**Figure 6:**
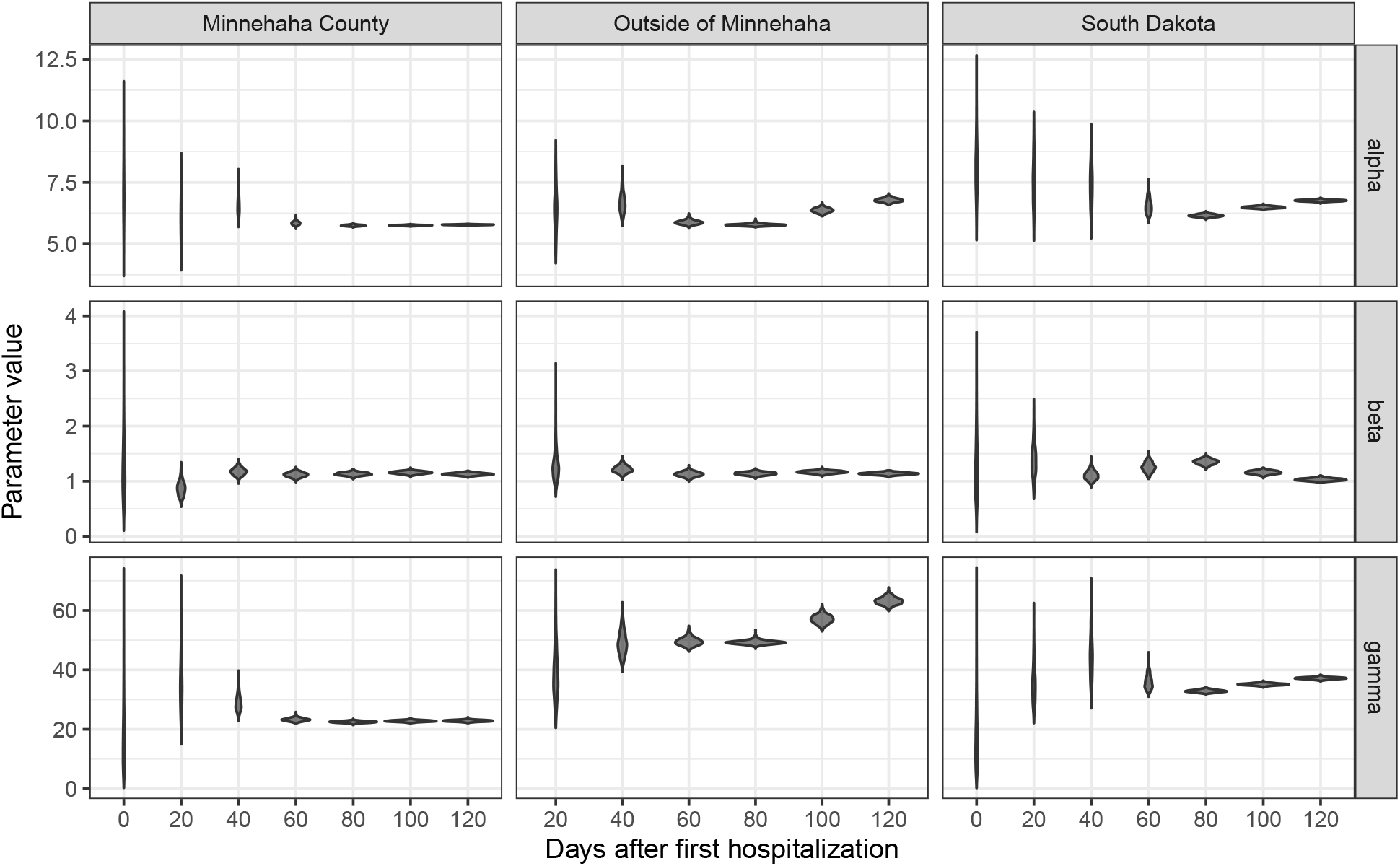
Change in parameter values over time. Violins represent posterior distributions of parameter values over time as models are fit using data at day 0, 20, 40, 60, 80, 100, and the most recent date.

## 12 Trace Plots

*Model 1*

\## [[1]]

**Figure 7:**
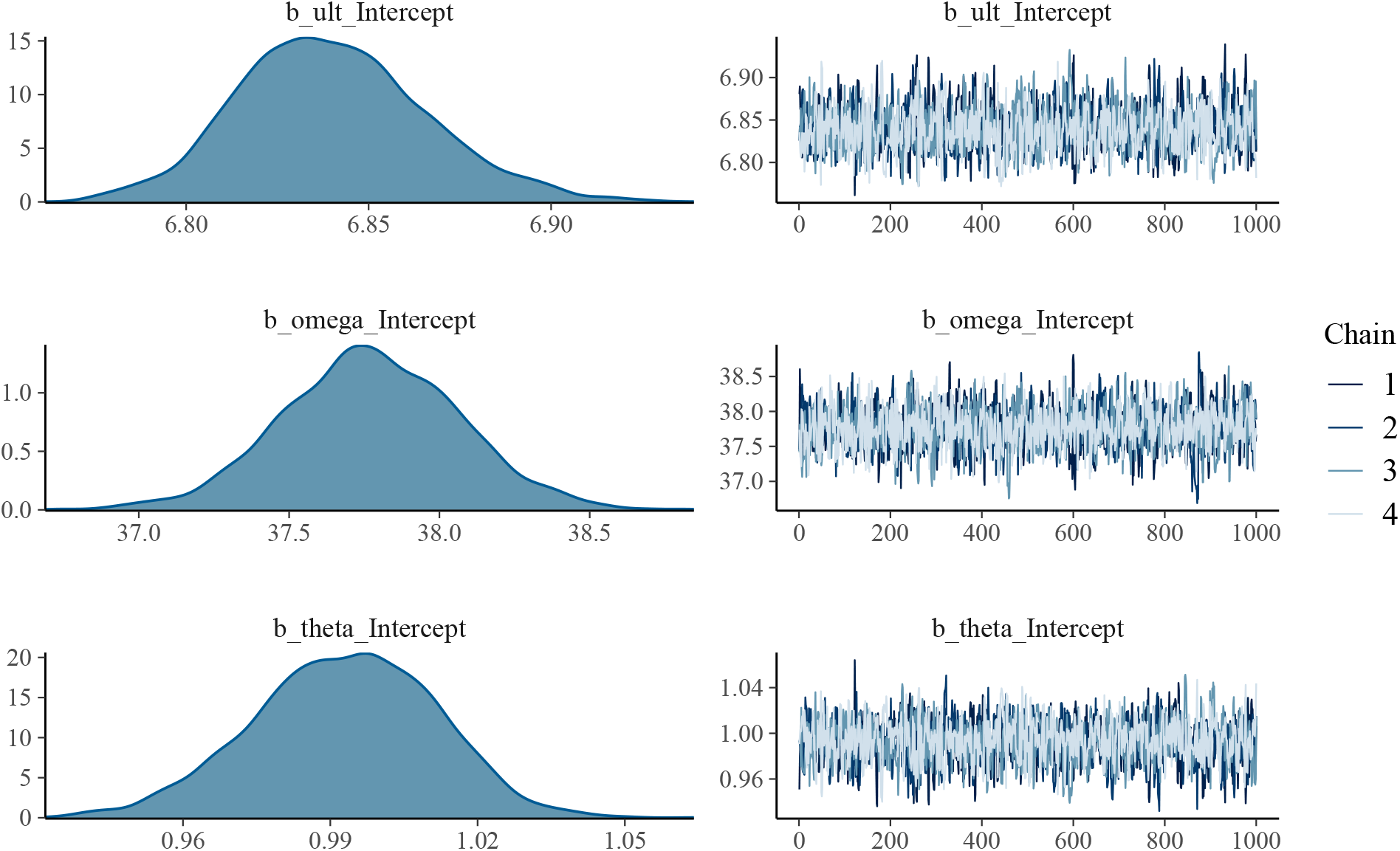
Posterior distributions and trace plots for each parameter from Model 1.

*Model 2*

\## [[1]]

**Figure 8:**
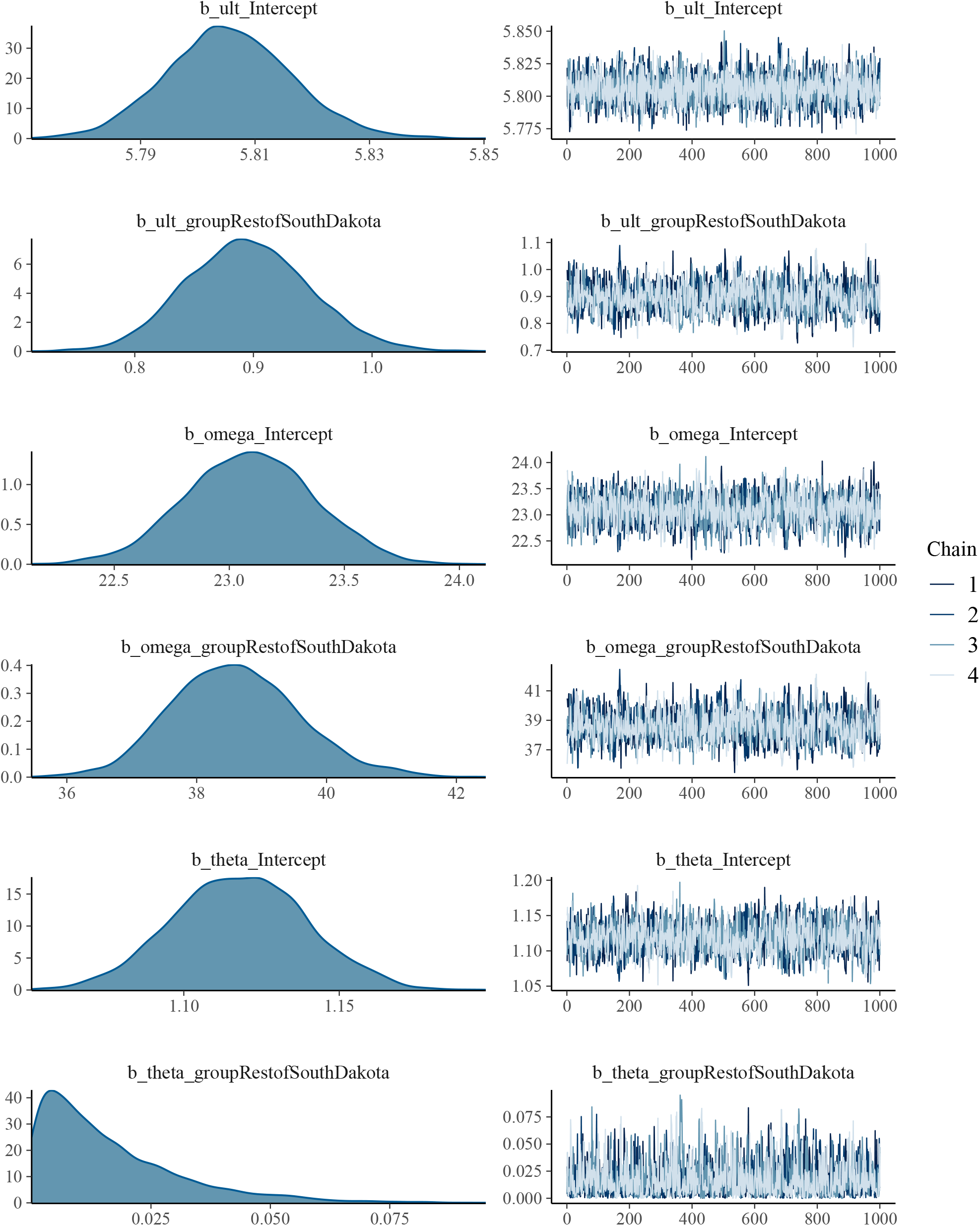
Posterior distributions and trace plots for each parameter from Model 2.

## 13 Stan code

Model code in the Stan language is below.

*Model 1*

~~~
## // generated with brms 2.12.0 ## functions {
## }
## data {
##  int<lower=1> N; // number of observations ##  int Y[N]; // response variable
##  int<lower=1> K_ult; // number of population-level effects
##  matrix[N, K_ult] X_ult; // population-level design matrix
##  int<lower=1> K_omega; // number of population-level effects
##  matrix[N, K_omega] X_omega; // population-level design matrix
##  int<lower=1> K_theta; // number of population-level effects
##  matrix[N, K_theta] X_theta; // population-level design matrix
##  // covariate vectors for non-linear functions
##  vector[N] C_1;
##  int prior_only; // should the likelihood be ignored?
## }
## transformed data {
## }
## parameters {
##  vector[K_ult] b_ult; // population-level effects
##  vector[K_omega] b_omega; // population-level effects
##  vector[K_theta] b_theta; // population-level effects
## }
## transformed parameters {
## }
## model {
##  // initialize linear predictor term
##  vector[N] nlp_ult = X_ult * b_ult;
##  // initialize linear predictor term
##  vector[N] nlp_omega = X_omega * b_omega;
##  // initialize linear predictor term
##  vector[N] nlp_theta = X_theta * b_theta;
##  // initialize non-linear predictor term
##  vector[N] mu;
##  for (n in 1:N) {
##   // compute non-linear predictor values
##   mu[n] = nlp_ult[n] * (1 - exp(- (C_1[n] / nlp_omega[n]) ^ nlp_theta[n]));
##  }
##  // priors including all constants
##  target += gamma_lpdf(b_ult | 64, 8);
##  target += gamma_lpdf(b_omega | 2.9, 0.17);
##  target += gamma_lpdf(b_theta | 5.8, 4.8);
##  // likelihood including all constants
##  if (!prior_only) {
##   target += poisson_log_lpmf(Y | mu);
##  }
## }
## generated quantities {
## // additionally draw samples from priors
## real prior_b_ult = gamma_rng(64,8);
## real prior_b_omega = gamma_rng(2.9,0.17);
## real prior_b_theta = gamma_rng(5.8,4.8);
## }
~~~

*Model 2*

~~~
## // generated with brms 2.12.0
## functions {
## }
## data {
##  int<lower=1> N; // number of observations
##  int Y[N]; // response variable
##  int<lower=1> K_ult; // number of population-level effects
##  matrix[N, K_ult] X_ult; // population-level design matrix
##  int<lower=1> K_omega; // number of population-level effects
##  matrix[N, K_omega] X_omega; // population-level design matrix
##  int<lower=1> K_theta; // number of population-level effects
##  matrix[N, K_theta] X_theta; // population-level design matrix
##  // covariate vectors for non-linear functions
##  vector[N] C_1;
##  int prior_only; // should the likelihood be ignored?
## }
## transformed data {
## }
## parameters {
##  vector<lower=0>[K_ult] b_ult; // population-level effects
##  vector<lower=0>[K_omega] b_omega; // population-level effects
##  vector<lower=0>[K_theta] b_theta; // population-level effects
## }
## transformed parameters {
## }
## model {
##  // initialize linear predictor term
##  vector[N] nlp_ult = X_ult * b_ult;
##  // initialize linear predictor term
##  vector[N] nlp_omega = X_omega * b_omega;
##  // initialize linear predictor term
##  vector[N] nlp_theta = X_theta * b_theta;
##  // initialize non-linear predictor term
##  vector[N] mu;
##  for (n in 1:N) {
##  // compute non-linear predictor values
##  mu[n] = nlp_ult[n] * (1 - exp(- (C_1[n] / nlp_omega[n]) ^ nlp_theta[n]));
##  }
##  // priors including all constants
##  target += gamma_lpdf(b_ult[1] | 49, 7)
##  - 1 * gamma_lccdf(0 | 49, 7);
##  target += normal_lpdf(b_ult[2] | 0, 0.5)
##  - 1 * normal_lccdf(0 | 0, 0.5);
##  target += gamma_lpdf(b_omega[1] | 2.9, 0.17)
##  - 1 * gamma_lccdf(0 | 2.9, 0.17);
##  target += normal_lpdf(b_omega[2] | 0, 5)
##  - 1 * normal_lccdf(0 | 0, 5);
##  target += gamma_lpdf(b_theta[1] | 5.8, 4.8)
##  - 1 * gamma_lccdf(0 | 5.8, 4.8);
##  target += normal_lpdf(b_theta[2] | 0, 1)
##  - 1 * normal_lccdf(0 | 0, 1);
##  // likelihood including all constants
##  if (!prior_only) {
##  target += poisson_log_lpmf(Y | mu);
##  }
## }
## generated quantities {
##  // additionally draw samples from priors
##  real prior_b_ult_1 = gamma_rng(49,7);
##  real prior_b_ult_2 = normal_rng(0,0.5);
##  real prior_b_omega_1 = gamma_rng(2.9,0.17);
##  real prior_b_omega_2 = normal_rng(0,5);
##  real prior_b_theta_1 = gamma_rng(5.8,4.8);
##  real prior_b_theta_2 = normal_rng(0,1);
##  // use rejection sampling for truncated priors
##  while (prior_b_ult_1 < 0) {
##   prior_b_ult_1 = gamma_rng(49,7);
##  }
##  while (prior_b_ult_2 < 0) {
##   prior_b_ult_2 = normal_rng(0,0.5);
##  }
##  while (prior_b_omega_1 < 0) {
##   prior_b_omega_1 = gamma_rng(2.9,0.17);
##  }
##  while (prior_b_omega_2 < 0) {
##   prior_b_omega_2 = normal_rng(0,5);
##  }
##  while (prior_b_theta_1 < 0) {
##   prior_b_theta_1 = gamma_rng(5.8,4.8);
##  }
##  while (prior_b_theta_2 < 0) {
##   prior_b_theta_2 = normal_rng(0,1);
##  }
## }
~~~

